# Effect of asymptomatic transmission and emergence time on multi-strain viral disease severity

**DOI:** 10.1101/2021.04.18.21255684

**Authors:** A. R. Alizad-Rahvar, M. Sadeghi

## Abstract

**Background:** In a viral epidemic, the emergence of a novel strain with increased transmissibility (larger value of basic reproduction number *R*_0_) sparks the fear that the increase in transmissibility is likely to lead to an increase in disease severity. It is required to investigate if a new, more contagious strain will be necessarily dominant in the population and resulting in more disease severity.

**Methods:** The impact of the asymptomatic transmission and the emergence time of a more transmissible variant of a multi-strain viral disease on the disease prevalence, disease severity, and the dominant variant in an epidemic was investigated by a proposed 2-strain epidemic model, called 2-SEICARD model, that is an extension of the SEIRD model.

**Results:** The simulation results showed that considering only *R*_0_, is insufficient to predict the outcome of a new, more contagious strain in the population. A more transmissible strain with a high fraction of asymptomatic cases can substantially reduce the mortality rate. If the emergence time of the new strain is closer to the start of the epidemic, the new, more contagious variant has more chance to win the viral competition and be the dominant strain; otherwise, despite being more contagious, it cannot dominate previous strains.

**Conclusions:** Three factors of *R*_0_, the fraction of asymptomatic transmission, and the emergence time of the new strain are required to correctly determine the prevalence, disease severity, and the winner of the viral competition.

## I. Introduction

The emergence of the novel coronavirus strain in the UK, called SARS-CoV-2 VOC 202012/01 or B.1.1.7, was shocking because this novel variant could be up to about 70% more transmissible than pre-existing variants of SARS-CoV-2^1^. This increased transmissibility can add between 0.4 and 0.7 to the basic reproduction number R_0_. This news sparked the fear that the increase in transmissibility is likely to lead to a large increase in hospitalization, intensive care unit (ICU) admission rate, and mortality. However, the studies about previous variants of SARS-CoV-2 showed that despite the rise of the lab-confirmed cases, the COVID-19 case fatality rate (CFR) declined, i.e., more transmissibility did not necessarily cause more severity^2,3^. Other studies in the UK and England showed that besides increasing the COVID-19 cases, the hospitalization rate, the ICU admission rate, and the CFR declined^4,5^. The preliminary explanation was the predominant shift towards positivity in younger age groups who have a better outcome. However, the analysis of German COVID-19 data^6^, which was reported by age categories, showed that the COVID-19 CFR declined across all age groups^7^. Interestingly, the older groups drove the overall reduction in CFR.

The public health authorities need to pinpoint the cause of this decline in the fatality rate in order to decide how to react against the newly emerged viral strains. The decision to fight blindly against a novel strain because of its increased transmissibility is not necessarily the most comprehensive and effective solution. We need to take other factors along with the transmissibility into account in our decision-making.

The spread of COVID-19 is an iceberg with the invisible part of being the asymptomatic transmission. The percent of asymptomatic cases who never experience COVID-19 symptoms remains uncertain. From about 20% to 50% of infected people are reported to be asymptomatic^8–10^. In a study, 39% of children aged 6-13 years tested positive for COVID-19 with no symptoms^11^. Different studies reported an insignificant difference in the upper respiratory viral load between symptomatic and asymptomatic cases^8,12^. Even a new study found that asymptomatic patients had higher SARS-CoV-2 viral loads than symptomatic cases^13^. Consequently, the asymptomatic infected people could play a significant driver role in the community spread of COVID-19. The results of a study demonstrated that both *R*_0_ and the proportion of asymptomatic transmissions were the main factors in controlling an infectious disease outbreak^14^.

In this study, we investigate the effect of the emergent viral strain on the total number of infected people and the illness severity by using epidemiological modeling. We will show that in an epidemic situation, the emergence time of the new strain and the relative *R*_0_ of the primary and the emergent strains determine the winner of the competition between two viral strains. Moreover, we will see that the disease severity and the mortality rate can be significantly influenced by the emergence time and the fraction of asymptomatic infectious cases of the emergent strain.

## II. Methods

For each viral strain, we use the extended version of the classic SEIRD epidemic model, called the SEICARD model, consisting of susceptible (S), exposed (in the latent period) (E), symptomatic infected (I), critically infected (C), asymptomatic infected (A), recovered (R), and dead (D) people. By paralleling two SEICARD models, we develop a 2-strain model, called 2-SEICARD, that describes the existence and competition of two viral variants in the population (Fig. 1). The index *s* = 1 or 2 in *E*_*s*_, *I*_*s*_, *C*_*s*_, *A*_*s*_, *R*_*s*_, and *D*_*s*_ represents the infectious strain in each group. It is assumed that the emergence time of the second strain is *T*_*E*_ days after the emergence time of the primary one, which is day 0. Moreover, we assume that there is no viral superinfection, i.e., the reinfection or co-infection between variants does not occur. In other words, the recovered individuals are cross-immunized and are immune to the new variants.

**Fig. 1.**
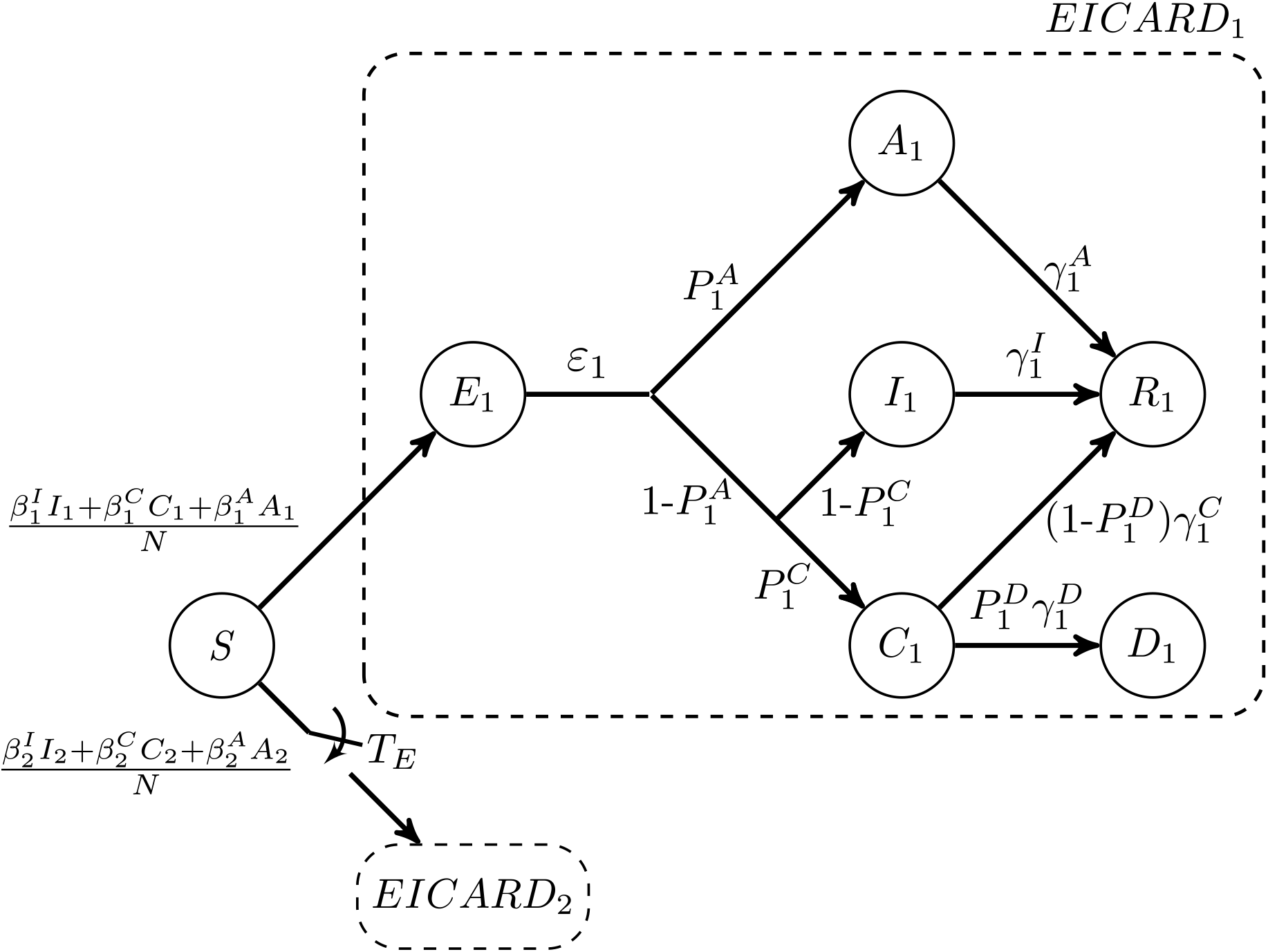
Schematic of the 2-SEICARD model. For simplicity, the lower branch, corresponding to the second strain, is not depicted, and it is denoted by *EICARD*_2_. The *EICARD*_2_ branch appears in the model for *t* ⩾ *T*_*E*_.

The parameters of the 2-SEICARD model are explained in Table I. The ODE system of this 2-strain model is given by

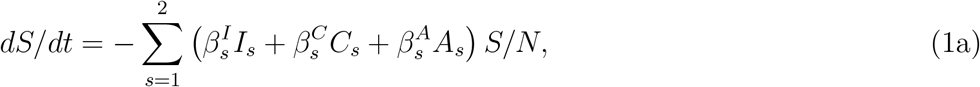

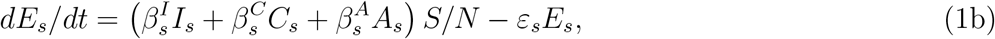

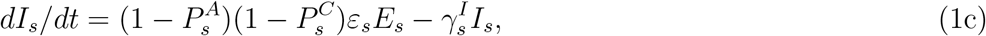

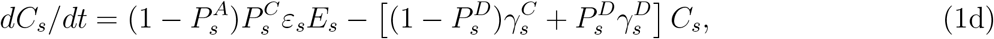

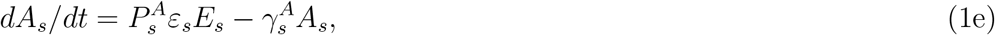

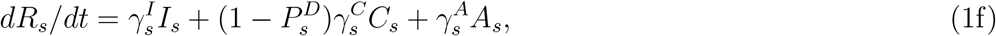

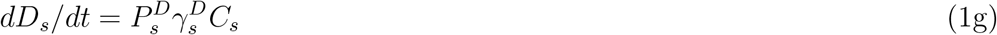

for *s* = 1 and 2. Here, the total population is 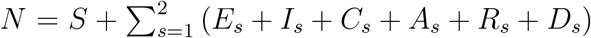. For simplicity, the natural birth and death rates are ignored in the model. To implement the emergence time *T*_*E*_, we set all parameters of the second strain to zero for *t* < *T*_*E*_.

**TABLE I.** Explanation of the symbols of the 2-SEICARD model.

| Symbol | Explanation | Value |
| --- | --- | --- |
| $s$ | Strain number | 1, 2 |
| $S$ | Susceptible individuals | |
| $E_s$ | Exposed to strain $s$ and still in the latent period | |
| $I_s$ | Symptomatic individuals infected with strain $s$ | |
| $C_s$ | Critically infectious individuals infected with strain $s$ | |
| $A_s$ | Asymptomatic individuals infected with strain $s$ | |
| $R_s$ | Individuals recovered from strain $s$' infection | |
| $D_s$ | Dead individuals infected with strain $s$ | |
| $N$ | Total number of individuals | 10000 |
| $P_s^A$ | Fraction of asymptomatic individuals infected with strain $s$ | 0.1 ($s = 1$)<br>0.1, 0.2, 0.4 ($s = 2$) |
| $P_s^C$ | Fraction of symptomatic individuals who are critically infected with strain $s$ | 0.1 |
| $P_s^D$ | Fraction of critically infected individuals who die from infection with strain $s$ | 0.05 |
| $\beta_s^I, \beta_s^C, \beta_s^A$ | Infection rate of different outcomes of strain $s$ | 0.2 ($s = 1$)<br>0.13, 0.2, 0.27 ($s = 2$) |
| $1/\varepsilon_s$ | Average incubation period of strain $s$ | 5 days |
| $1/\gamma_s^I, 1/\gamma_s^C, 1/\gamma_s^A, 1/\gamma_s^D$ | Average infection period of different outcomes of strain $s$ | 10 days |
| $T_E$ | Emergence time of strain 2 | $0 \leq T_E \leq 100$ day |

For each strain, the infection rates 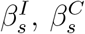, and 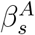 denote the probability of transmitting disease from *I*_*s*_, *C*_*s*_, or *A*_*s*_ to *S*, respectively. On the other hand, as Fig. 1 shows, the outcome of each exposed person *E*_*s*_ could be *A*_*s*_, *I*_*s*_, *C*_*s*_, and *D*_*s*_ with the probabilities of 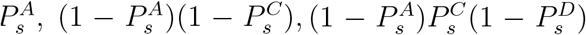, and 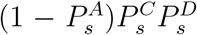, respectively. By using the method of next-generation matrices^15^, we can obtain the following expression for the *R*_0_ of each strain

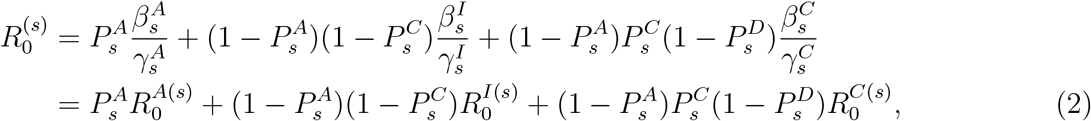

where 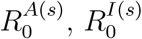, and 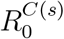 denote the reproduction number of each outcome, and 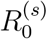 is obtained by their weighted sum. The weight of each outcome is the probability of its occurrence.

The values of different parameters used in the 2-SEICARD model are listed in Table I. In our modeling, we assume a wild animal population in which there is no isolation and social policy or restriction. Hence, we consider that all *I*_*s*_, *C*_*s*_, and *A*_*s*_ outcomes have the same probability of transmission; i.e., 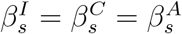. According to Eq. (2), by considering 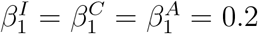 and the values of 0.13, 0.2, and 0.27 for all *β* values of strain 2, we have 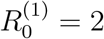 and 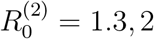, and 2.7, respectively.

## III. Results and Discussion

In this section, we consider fixed parameters for the first strain in the 2-SEICARD model; i.e., 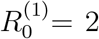 and 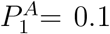. Then, the emergent strain with different values of 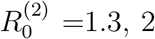, and 2.7 and 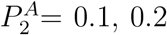 and 0.4 emerges at day *T*_*E*_, where 0 ⩽ *T*_*E*_ ⩽ 100. In all the above scenarios, we study the effect of the emergent strain on the total number of infected cases and the mortality rate, as a measure of severity, from the beginning of the epidemic until we reach the endemic steady state. The total number of infected cases is *N* − *S*_∞_, where *S*_∞_ denotes the number of susceptible cases that have not been infected at all when the disease has gone. The mortality rate is the total proportion of deaths in the population due to infection.

### A. Effect of R_0_ and T_E_ on the total number of infections and the dominant strain

The simulation results show that the total number of infected cases does not vary with 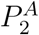 for the fixed values of 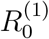 and 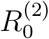. In other words, the values of 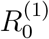 and 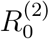 determine the total number of infected individuals during the epidemic spread. Provided that 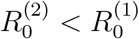, the emergent strain does not have any chance to compete with the primary strain and would become extinct immediately (see Fig. 2(A1)). In the case of 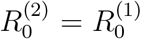, the total number of infected cases with two strains remains the same as that in the case of spreading only the primary strain in the population with the same value of the basic reproduction number. Moreover, Fig. 2(A2) demonstrates that the total number of infected cases does not vary with the emergence time of the second strain, *T*_*E*_. However, the later emergence of strain 2 results in less proportion of infection with this strain in the population. In contrast, Fig. 2(A3) depicts that the emergence of a more contagious strain 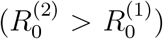 increases the total number of infected cases compared to the existence of only the primary strain. Furthermore, Fig. 2(A3) shows that the new, more contagious strain with larger value of *R*_0_ does not necessarily dominate in the population. In other words, *R*_0_ alone does not determine which virus wins the viral competition. Indeed, besides the values of *R*_0_, the emergence time *T*_*E*_ also determines whether the new strain with more transmissibility dominates the primary one or not. The sooner emergence of the new, more contagious variant can make it dominant; otherwise, the primary strain remains dominant in the population.

**Fig. 2.**
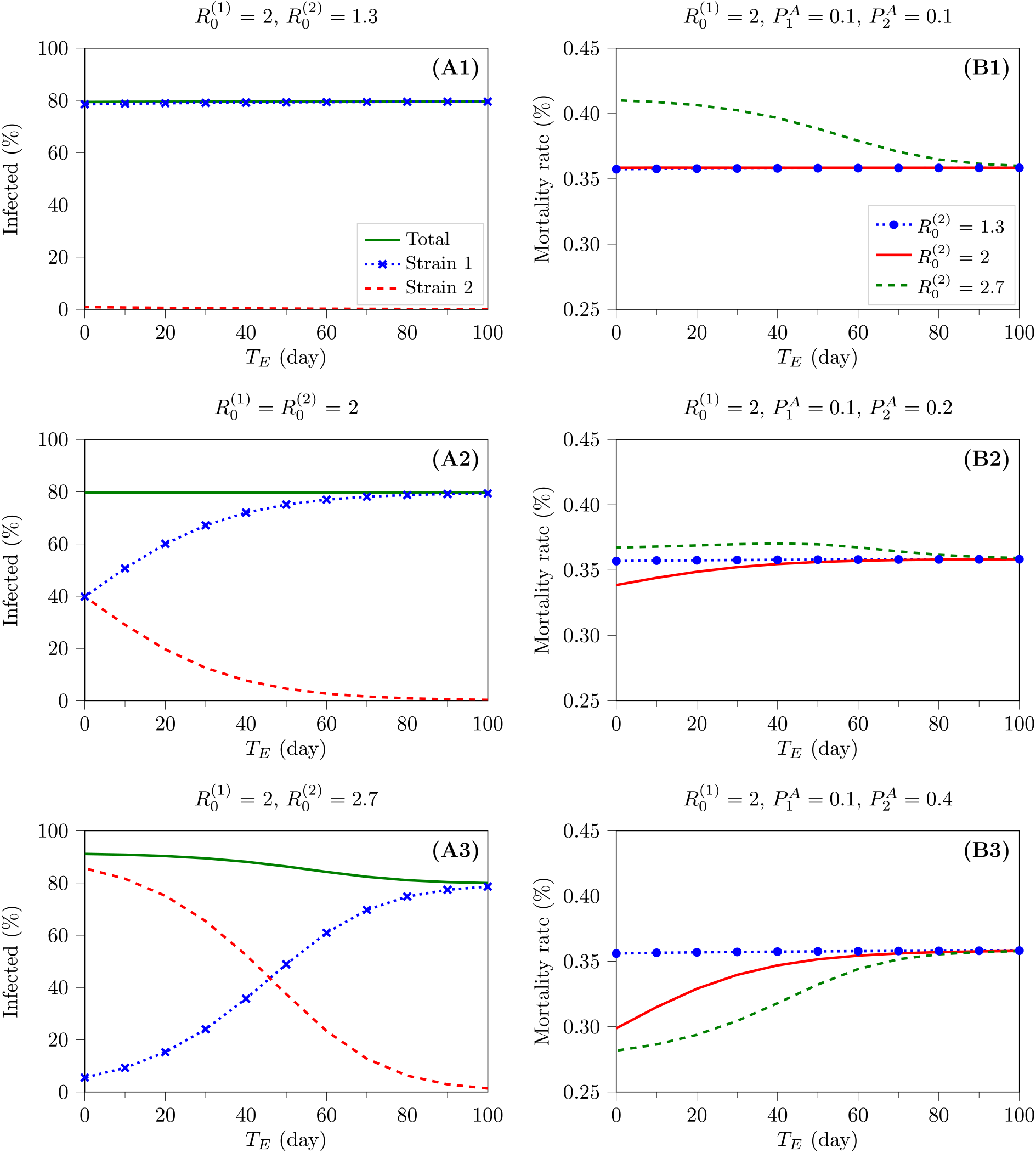
The effect of 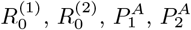, and *T*_*E*_ on the total number of infected cases (A1-A3) and the mortality rate (B1-B3). These figures indicate that increased transmissibility of the new emergent strain does not necessarily reflect more severity of the disease. **A1-A3:** The fraction of the population who are infected with strains 1 and 2 and the total percentage of the infected cases are depicted for three cases of 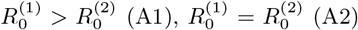, and 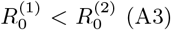. Also, these figures demonstrate that the emergent time of the new strain (*T*_*E*_) should be considered to determine the winner of the viral competition. The legends of all figures are the same as those in Fig. A1. **B1-B3**: For 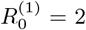 and 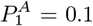, these figures show the effect of increase in the fraction of asymptomatic cases of the emergent strain (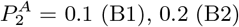, and 0.4 (B3)), on the mortality rate for different levels of transmissibility of the new variant (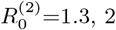, and 2.7). The legends of all figures are the same as those in Fig. B1.

### B. Effect of emergent strain on mortality rate

Our main question is that in the case of 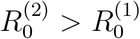, whether or not more infected cases increase similarly the mortality rate of the disease or not. To answer this question, consider the mortality rate in different circumstances in Figures 2(B1-B3). These figures show concurrently the effect of the fraction of asymptomatic individuals infected with strain 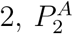, and the emergence time of the second strain, *T*_*E*_, on the mortality rate for 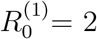 and 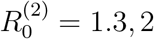, and 2.7. As we have discussed, if 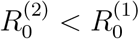, the emergent strain cannot compete with the primary strain, and hence, the mortality rate remains fixed for all values of 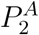, equal to the mortality rate of the primary strain alone. Hence, in these figures, although blue curves are corresponding to the 2-strain scenario, they also represent the mortality rate of the primary strain alone.

As we expected, provided that 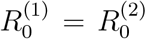 and 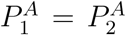, the mortality rate remains the same as that of the primary strain alone. On the other hand, as Fig. 2(B1) depicts, if both strains have a similar proportion of asymptomatic cases, i.e., 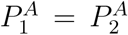, the mortality rate increases with the emergence of a more contagious strain 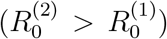. In this case, the sooner that the new strain emerges, the more in the mortality rate increases. However, as Figures 2(B2-B3) show, the increase in 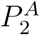 can reduce the mortality rate for 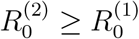. For large values of 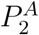, although a more contagious strain emerges, it can make the mortality rate less than that before the emergence (Fig. 2(B3)). Interestingly, for large values of 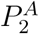, the emergent strain with higher *R*_0_ decreases the mortality rate more. In other words, more transmissibility does not reflect necessarily more severity, and both 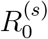 and 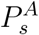 values should be considered to correctly determine the effect of the more transmissible new strain on the viral disease severity.

## IV. Conclusion

In this study, we investigated the impact of the asymptomatic transmission and the emergence time of the new, more contagious viral strain on the disease prevalence, disease severity, and the dominant variant in an epidemic. Our results demonstrated that being an emergent strain with more transmissibility, i.e., having a larger basic reproduction number *R*_0_, compared to previous variants, does not necessarily lead to more severity or mean that the new variant will dominate in the population. Indeed, when a new strain with a larger basic reproduction number emerges, it will increase the number of infected cases in the population. However, the creation of more severe outcomes depends on the fraction of asymptomatic transmissions. If a large proportion of infections, due to the new variant, do not show any symptom, they can even reduce the mortality rate in the population. Moreover, provided that the emergence time of the new strain is closer to the start of the epidemic, the new, more contagious variant has more chance to win the viral competition and be the dominant strain; otherwise, despite being more contagious, it cannot dominate previous strains.

## Data Availability

No data is available.

## Acknowledgement

The authors would like to thank A. Kalirad for constructive discussion that contributed to the development of this research.

## References

[1] Davies NG, Barnard RC, Jarvis CI, Kucharski AJ, Munday J, Pearson CAB, et al. Estimated transmissibility and severity of novel SARS-CoV-2 Variant of Concern 202012/01 in England. medRxiv. 2020.

[2] SARS-CoV-2 Variants;. Available from: www.who.int/csr/don/31-december-2020-sars-cov2-variants/.

[3] Why do COVID death rates seem to be falling?;. Available from: www.nature.com/articles/d41586-020-03132-4.

[4] Howdon D, Oke J, Heneghan C. COVID-19: Declining Admissions to Intensive Care Units;. Available from: www.cebm.net/covid-19/covid-19-declining-admissions-to-intensive-care-units.

[5] Howdon D, Heneghan C. The Declining Case Fatality Ratio in England;. Available from: www.cebm.net/covid-19/the-declining-case-fatality-ratio-in-england.

[6] RKI COVID19;. Available from: https://npgeo-corona-npgeo-de.hub.arcgis.com/datasets/dd4580c810204019a7b8eb3e0b329dd60/data.

[7] Oke J, Howdon D, Heneghan C. Declining COVID-19 Case Fatality Rates across all ages: analysis of German data;. Available from: https://www.cebm.net/covid-19/declining-covid-19-case-fatality-rates-across-all-ages-analysis-of-german-data.

[8] Ra SH, Lim JS, Kim Gu, Kim MJ, Jung J, Kim SH. Upper respiratory viral load in asymptomatic individuals and mildly symptomatic patients with SARS-CoV-2 infection. Thorax. 2021;76(1):61–63.

[9] Gudbjartsson DF, Helgason A, Jonsson H, Magnusson OT, Melsted P, Norddahl GL, et al. Spread of SARS-CoV-2 in the Icelandic Population. New England Journal of Medicine. 2020;382(24):2302–2315.

[10] Byambasuren O, Cardona M, Bell K, Clark J, McLaws ML, Glasziou P. Estimating the extent of asymptomatic COVID-19 and its potential for community transmission: Systematic review and meta-analysis. Journal of the Association of Medical Microbiology and Infectious Disease Canada. 2020;5:223–234.

[11] Hurst JH, Heston SM, Chambers HN, Cunningham HM, Price MJ, Suarez L, et al. Severe Acute Respiratory Syndrome Coronavirus 2 Infections Among Children in the Biospecimens from Respiratory Virus-Exposed Kids (BRAVE Kids) Study. Clinical Infectious Diseases. 2020 11. Available from: https://doi.org/10.1093/cid/ciaa1693.

[12] Cereda D, Tirani M, Rovida F, Demicheli V, Ajelli M, Poletti P, et al. The early phase of the COVID-19 outbreak in Lombardy, Italy. arXiv. 2020. Available from: https://arxiv.org/abs/2003.09320.

[13] Hasanoglu I, Korukluoglu G, Asilturk D, Cosgun Y, Kalem AK, Altas AB, et al. Higher viral loads in asymptomatic COVID-19 patients might be the invisible part of the iceberg. Infection. 2020 Nov.

[14] Fraser C, Riley S, Anderson RM, Ferguson NM. Factors that make an infectious disease outbreak controllable. Proceedings of the National Academy of Sciences. 2004;101(16):6146–6151. Available from: https://www.pnas.org/content/101/16/6146.

[15] Diekmann O, Heesterbeek JA, Roberts MG. The construction of next-generation matrices for compartmental epidemic models. J R Soc Interface. 2010 Jun;7(47):873–885.

